# Exploring regression dilution bias using repeat measurements of 2858 variables in up to 49 000 UK Biobank participants

**DOI:** 10.1101/2022.07.13.22277605

**Authors:** Charlotte E Rutter, Louise AC Millard, Maria Carolina Borges, Deborah A Lawlor

## Abstract

**Background:** Measurement error in exposures and confounders can bias exposure-outcome associations but is rarely considered. Our aim was to assess measurement error between repeat measures of all continuous variables in UK Biobank, and explore approaches to mitigate its impact on exposure-outcome associations.

**Methods:** Intraclass correlation coefficients (ICC) were calculated for all continuous variables with repeat measures. Regression calibration was used to correct for measurement error in both exposures and confounders, using the association of C-Reactive Protein (CRP) with mortality as an illustrative example.

**Results:** The 2858 continuous variables with repeat measures, varied in sample size from 109 to 49 121. They fell into three groups: (i) baseline visit measures (529 variables; median ICC=0.64, IQR= [0.57, 0.83]); (ii) online dietary measures (22 variables; median ICC=0.35, IQR=[0.30, 0.40]) and (iii) imaging measures (2307 variables; median ICC=0.85, IQR=[0.73, 0.94]). Highest ICC were for anthropometric and medical history measures, and lowest for dietary and heart magnetic resonance imaging.

The ICC for CRP was 0.29 (95% CI=[0.27, 0.30]), and for body mass index and smoking pack-years (confounders), were 0.93 (95% CI=[0.92, 0.93]) and 0.85 (95% CI=[0.84, 0.86]) respectively. The association of CRP with all-cause mortality (Hazard Ratio (HR)=1.029 per mg/L, 95% CI=[1.028, 1.031]) increased when correction for RDB was applied to the exposure (HR=1.119, 95% CI=[1.095, 1.143]). Confounder correction did not influence estimates.

**Conclusions:** Measurement error varies widely and is often non-negligible. For UK Biobank we provide relevant statistics and adaptable code to help other researchers explore and correct for bias due to measurement error.

**Key Messages:**

- Random measurement error in the exposure and confounders can bias the association between exposure and outcome towards or away from the null.
- Some prospective studies, including UK Biobank, provide repeat measures of variables in a sub-sample for exploring bias due to random measurement error; these are rarely used.
- Our results demonstrate that measurement error is often non-negligible and may bias estimates.
- Agreement between repeated measures varies by category. In UK Biobank, dietary measures and heart magnetic resonance imaging are least stable while anthropometric and medical history variables are most stable.
- We have provided intraclass correlation coefficients for 2858 continuous variables from UK Biobank and adaptable code to support researchers to correct for bias due to random error in exposures and confounders.

## Introduction

Measurement error is a widespread problem and can be random (i.e. where the errors in measurement average to zero) or systematic (where a measurement is on average higher or lower than the true value)^1,2^. Our focus is on random error where an underlying mean value of a variable is measured with random variation (either due to true fluctuations or imprecision of measurement).

Regression dilution bias (RDB), also known as attenuation by errors^3^, refers to attenuation of an estimate between any covariable (i.e. exposure, confounder or mediator) and an outcome toward the null, due to non-differential random measurement error in that covariable^3^ (non-differential meaning the error is not related to the outcome). Epidemiological studies often comment, in the discussion, on how findings might be underestimated because of RDB in an exposure, without considering random measurement error of confounders that may impact exposure estimates in unanticipated ways.^1^ It has also been noted that where random measurement error is controlled for, this is often only done for exposure-outcome associations without making any attempt to control for random error in confounders.^3,4^ Whilst each confounder-outcome association would be biased towards the null in the presence of random error in that confounder, the impact of this random error on the main exposure-outcome effect would depend on the (unbiased) direction and size of effect of the confounders on the exposure and outcome.^1^ Thus RDB in confounders, as well as in the exposure, could alter the exposure-outcome effect estimates in either direction.

Some studies invite a subset of participants back for repeat assessments with the express purpose of providing researchers with data that could be used to correct for random measurement error but these data seem to be rarely used. The Avon Longitudinal Study of Parents and Children (ALSPAC) repeated all 13 clinic assessments of parents and children, undertaken over the last ∼30 years^5,6^ in a subset of 3% of participants, and UK Biobank (UKB) did the same in up to ∼49 000 participants (∼10%) to enable assessment of potential random measurement error^7^. We have only identified three papers in ALSPAC^8-10^ and four papers in UKB^11-14^ that have utilised repeat measures; in all cases to correct for RDB in exposure-outcome associations only. Thus, even where data are available to address random measurement error it is rarely used and the research resources and participant time and effort to collect these data are largely wasted.

The aim of this paper was to assess measurement error between repeat measures of all continuous variables in UKB, and explore approaches to mitigate its impact when estimating exposure-outcome associations. First, we assessed random and systematic error using intraclass correlation coefficients (ICC)^16^ and accuracy coefficients^17, 18^, respectively. Second, we used an illustrative example to exemplify how to account for random error in exposure and confounders using ICCs and regression calibration. We provide ICCs and adaptable code describing how to adjust for RDB in exposure- and confounder-outcome associations, so that other researchers can assess and account for random measurement error in their own analyses. We have used UKB data as the scope for use was large; at the end of 2021, there were 25 000 registered researchers, with over 4600 publications, with the number increasing exponentially from 120 in 2016 to 1700 in 2021.^15^

## Methods

### Study data

UKB is a large prospective cohort study of over 500 000 adults aged 40-69 at recruitment (5.5% response of those invited), open to researchers for health-related research.^7^ The initial assessment, between 2006-10, included physical measurements, participant completed responses to computer questionnaires, as well as laboratory analysis of blood and urine samples. Around 20 000 participants were invited back for a repeat visit in 2012-13 when most measurements and sample assays were repeated. Some additional repeated information was collected via online questionnaires at other times for up to 49 000 participants, for example the 24-hour diet survey (here we use two repeat measures from comparable online surveys and not the measure from the baseline visit).

Imaging data was collected during visits starting in 2014 and repeated from 2019. Imaging data collection is on-going; here we present data for the first 4498 participants with repeat measures.

From the full list of variables available on UKB showcase^19^ we excluded those without repeat measures, those that were not feasible covariables in regression analyses (e.g. data processing information) and those that were not continuous measures (or integer measures that could be treated as continuous, having more than 20 distinct values). We further excluded variables where more than 20% of participants had the same value, and those where fewer than 100 participants had repeated measurements (Figure 1 and Supplementary Table S1).

**Figure 1.**
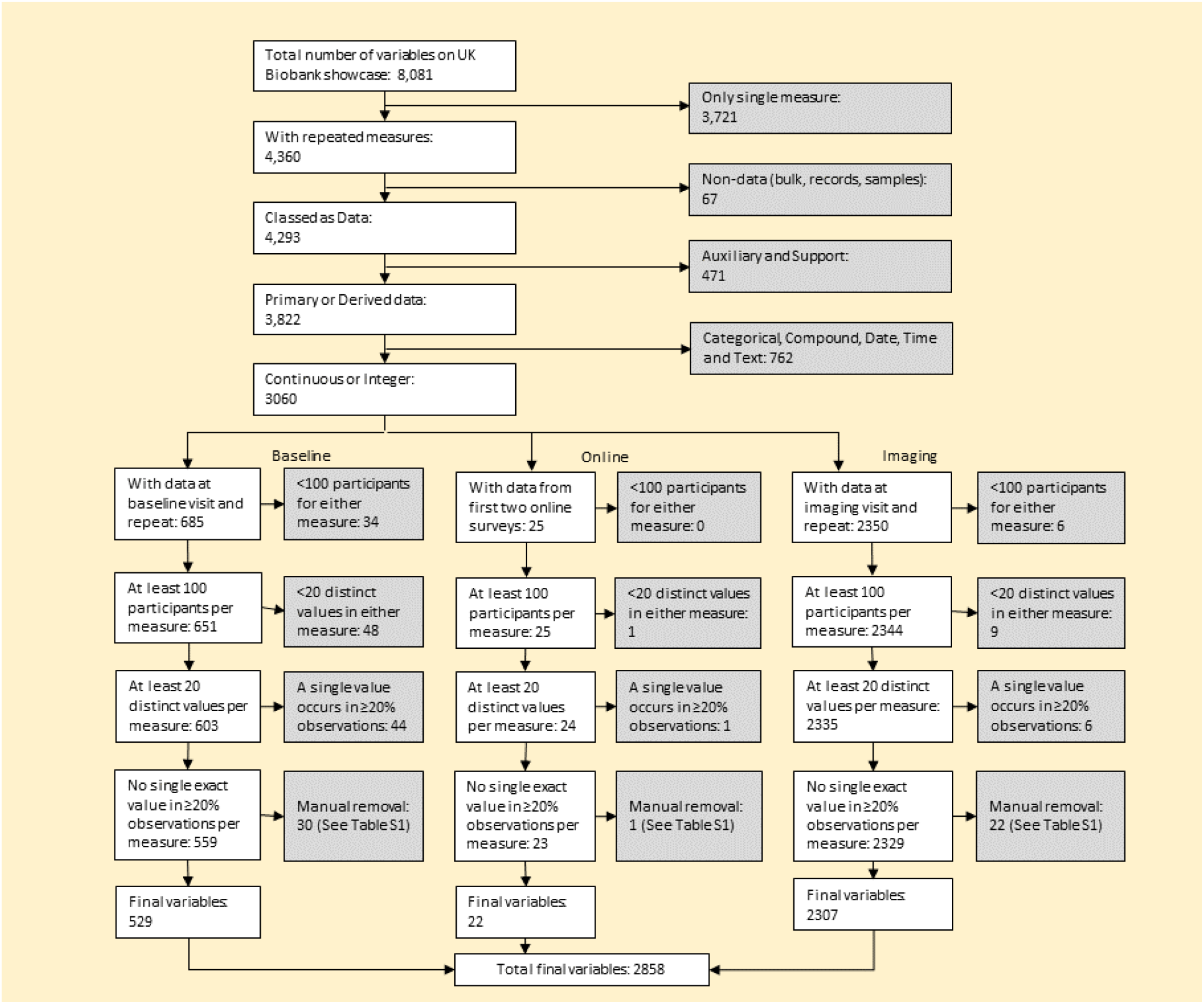
Data flow chart

Information on how we dealt with values expressed as e.g. ≤ 1mile and those with multiple readings at a visit is provided in Supplementary Text S1 and Tables S2 and S3.

### Statistical Analyses

#### Assessing error in repeat measures

We calculated the Intraclass Correlation Coefficient (ICC) for each variable, which shows the level of agreement between two sets of measurements. In this test-retest situation, i.e. repeated measures but separated by weeks or months and not necessarily under identical circumstances (e.g. the person doing the assessment may be different), the most appropriate method to assess the ICC uses a two-way mixed effects model^16^ (Supplementary Text S2). Higher ICC suggest lower random measurement error.

We also calculated the accuracy coefficient, a measure of systematic bias, using Lin’s method.^17,18^ The accuracy coefficient is a measure of how close the line of best fit between two repeat measures is to a line of 45 degrees through the origin (Supplementary Text S2). The higher the accuracy coefficient the less likely there is to be systematic error.

#### Correction for random measurement error

We focused on two commonly used methods: correction using the ICC,^20^ and regression calibration^21^. The ICC method can correct for RDB due to random error in the exposure, but it is not possible to account for random error in confounders using this method. It involves first fitting a regression model to estimate the exposure-outcome association, and then using the ICC statistic to correct this estimate and associated confidence interval.

The correction factor 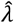 is the reciprocal of the ICC.^16^ The uncorrected main effect estimate 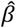 e.g. difference in means, log odds, logit, or log hazard is multiplied by the correction factor to get the corrected estimate 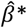. The confidence interval uses the following formula to take account of the uncertainty in the correction factor:

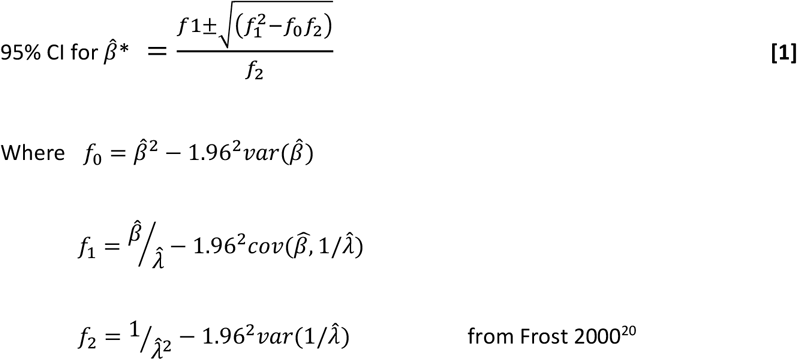

The covariance 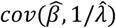 is zero if the sample of repeated measures is different to the main analysis sample. If the repeated sample is a small subset of the main sample then the covariance will be close to zero and the parameters can be treated as independent^20^, which is what we assume in this paper. Formulae for the variances of 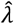 and 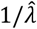 are in Supplementary Text S3.

Regression calibration can be used to correct for bias due to random error in the exposure and confounders (or other covariables) together. It involves regressing the repeat measure of the exposure on the initial measure, including in the model the same confounders used in the main analysis model [First stage model].^21^ Then the coefficients from this linear regression are used to predict fitted values of the exposure for the whole dataset. These predicted values are used in the main model in place of the original values of the exposure [Second stage model]. The effect estimate from this model is thus corrected for exposure-outcome RDB. To correct for random measurement error in both the confounders and exposure, a first stage model is run for each covariable measured with error, adjusted for all other covariables (e.g. exposure and confounders). Predicted values from each of these first stage models are then used in the second stage model. To account for uncertainty in both stages we recommend bootstrapping the whole process to obtain corrected confidence intervals.^21^

Both of the above methods can provide unbiased estimates of association with linear regression and approximately unbiased estimates for non-linear Cox proportional hazards and logistic regression.^21,22^

#### Illustrative example

Our illustrative example explores the association of C-Reactive Protein (CRP) with all-cause, cardiovascular and cancer mortality, using confounder adjusted Cox proportional hazards. CRP is a widely used biomarker of chronic inflammation. High values of CRP have been associated with increased risk of cardiovascular disease and colorectal, lung and breast cancers.^23-26^ In addition to these associations CRP is known to be highly variable across time.

Confounders were selected a priori, on the basis of being known or plausible determinants of both CRP and mortality. These were sex, age, ethnicity, home neighbourhood area deprivation score, lifetime smoking pack-years, alcohol consumption and BMI. Full details of how these were measured and categorised are provided in Supplementary Text S4. Four of these confounders – age, area deprivation, smoking and BMI – were continuous. Of these, repeat measures were not available for area deprivation. Difference in age between the two repeat measurements accurately reflected the participants aging and so was not considered a potential source of random measurement error. Thus, we corrected for potential bias due to random error in smoking and BMI.

We fitted Cox proportional hazards models using date of baseline assessment (i.e. when blood was drawn for CRP) and date of death or end of follow-up (28/02/2021) for those who survived, to calculate years of follow-up. Mortality data was available in the UK Biobank for all participants, via linkage to UK death registries.

For each outcome we present confounder-adjusted results with: (i) no correction for random measurement error; (ii) correction for RDB due to exposure random error using ICC; (iii) correction for RDB due to exposure random error using regression calibration; and (iv) correction for random error in both exposure and (continuously measured) confounders using regression calibration. Bootstrapping was used over the whole regression calibration process (first stage linear regression followed by second stage Cox proportional hazards) with 10 000 replicates, to calculate confidence intervals.

All code is available at https://github.com/MRCIEU/measurement_error_adjustment. Git tag v0.1 corresponds to the version presented here. All analysis was completed using Stata version 16.^27^

## Results

### Assessing error in repeat measures

Of the 8081 available UKB variables we were able to provide correction factors for 2858. Split into three groups, this included 529 baseline visit variables, 22 online diet questionnaire variables and 2307 medical imaging visit variables. Most excluded variables were due to not having a repeat measure (N = 3721) or not being continuously measured (N= 762). (Figure 1)

Supplementary tables S4-S6 and Supplementary figures S1-S10 provide metadata on the variables with repeat measures, including time between repeat measures, available sample size, ICC, accuracy coefficient, and correction factor statistics. The median time between the repeated visits in baseline measurements varied between 34 and 56 months. All of the online diet variables had a median of 4 months between visits and all of the imaging visit variables had a median of 27 months between visits. The sample size of repeated measures across all variables varied from 135 to 45 836.

Overall, the online diet variables had lowest ICC (median=0.35, IQR=[0.30, 0.40]), baseline visit variables in-between (median=0.64, IQR=[0.57,0.82]) and imaging visit variables the highest (median=0.85, IQR=[0.73, 0.94]). Across all variables, 7% had ICC <0.50, 28% had ICC between 0.50-0.74, 29% had ICC between 0.75-0.89 and 36% had ICC 0.90 or higher. By variable category, the highest ICC were seen in anthropometric, medical history and DXA scan variables and the lowest in dietary measures and heart MRI (Figure 2). Most variables had accuracy coefficients above 0.95 (Figure 3 and Supplementary tables S4-S6) indicating generally low systematic error, though the amount of systematic error differed by variable category (Figure 4 shows illustrative ICC and accuracy coefficients together for anthropometric, diet and heart MRI; with similar figures for individual variable categories in Supplementary figures S1-S10, except metabolomics, DXA scan and brain MRI due to number of variables). As an example, standing height had high ICC and accuracy, whereas seated height has high ICC and low accuracy (Figure 3). This is due to a change in the height of the box used to sit on between visits, which might not be realised by researchers without checking the repeat measurements and then the UKB documents to understand this.

**Figure 2.**
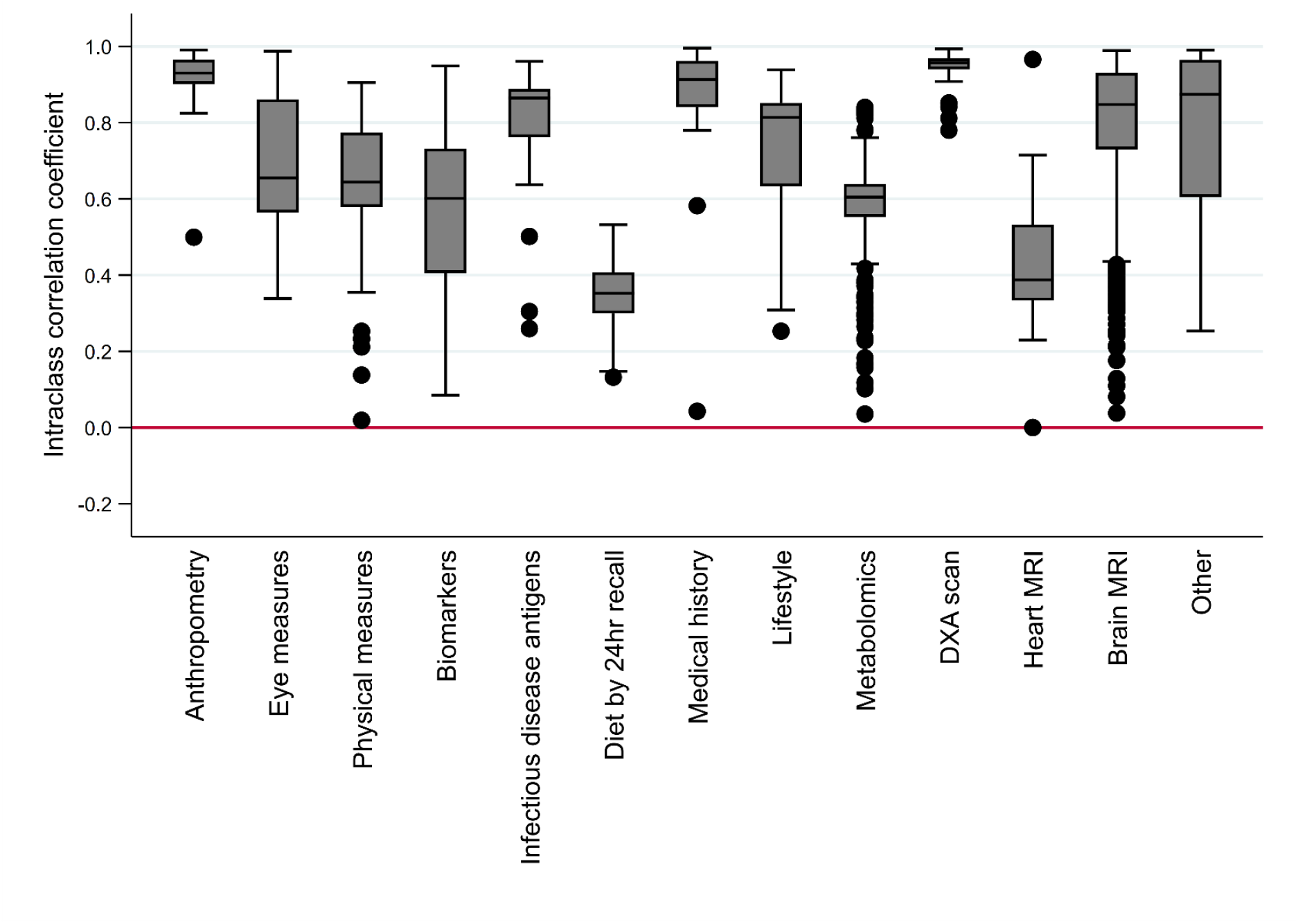
Intraclass correlation coefficient by variable category, measuring overall agreement between the initial and repeated measurements

**Figure 3.**
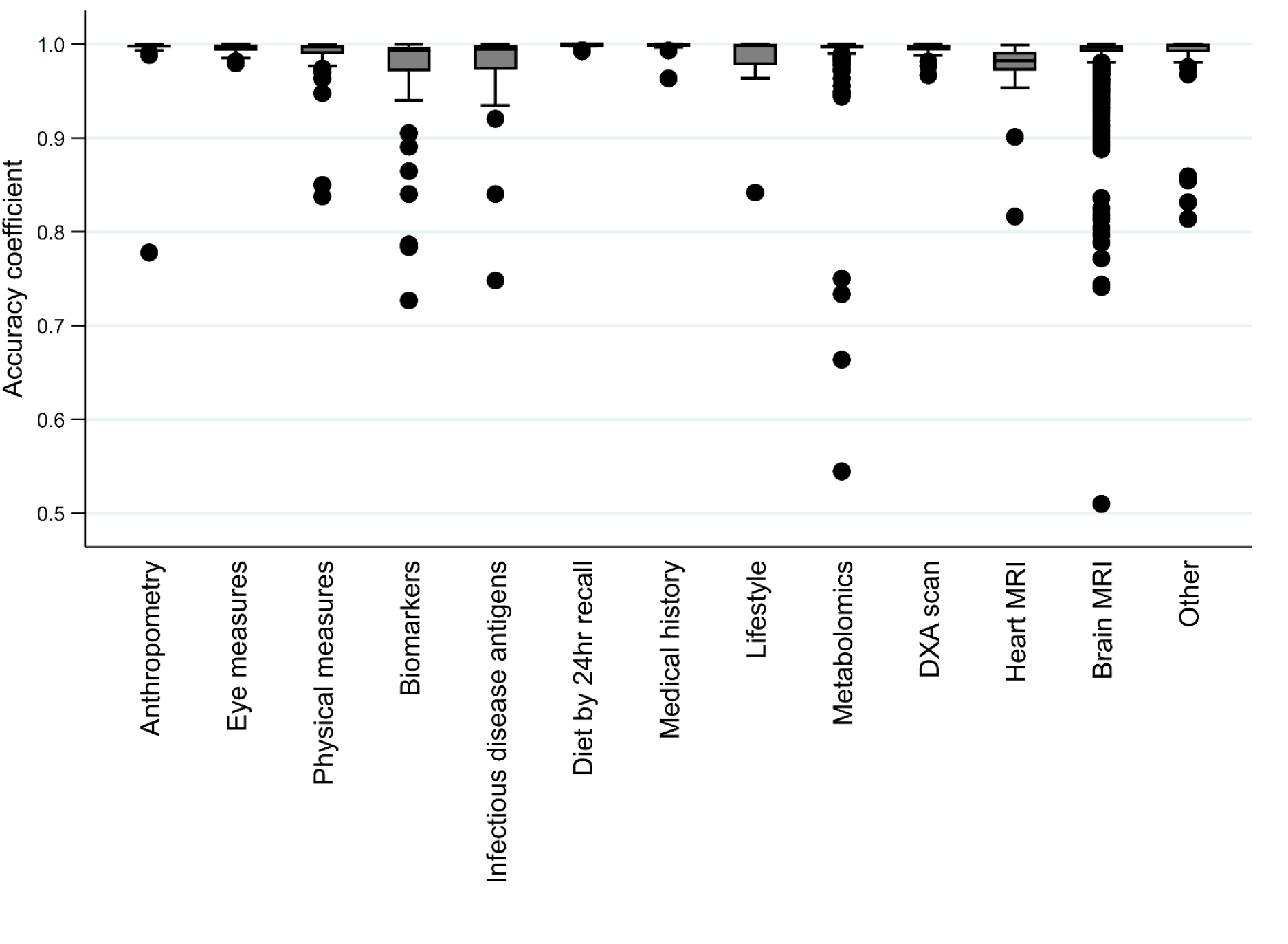
Accuracy coefficient by variable category, measuring systematic difference between the initial and repeated measurements

**Figure 4.**
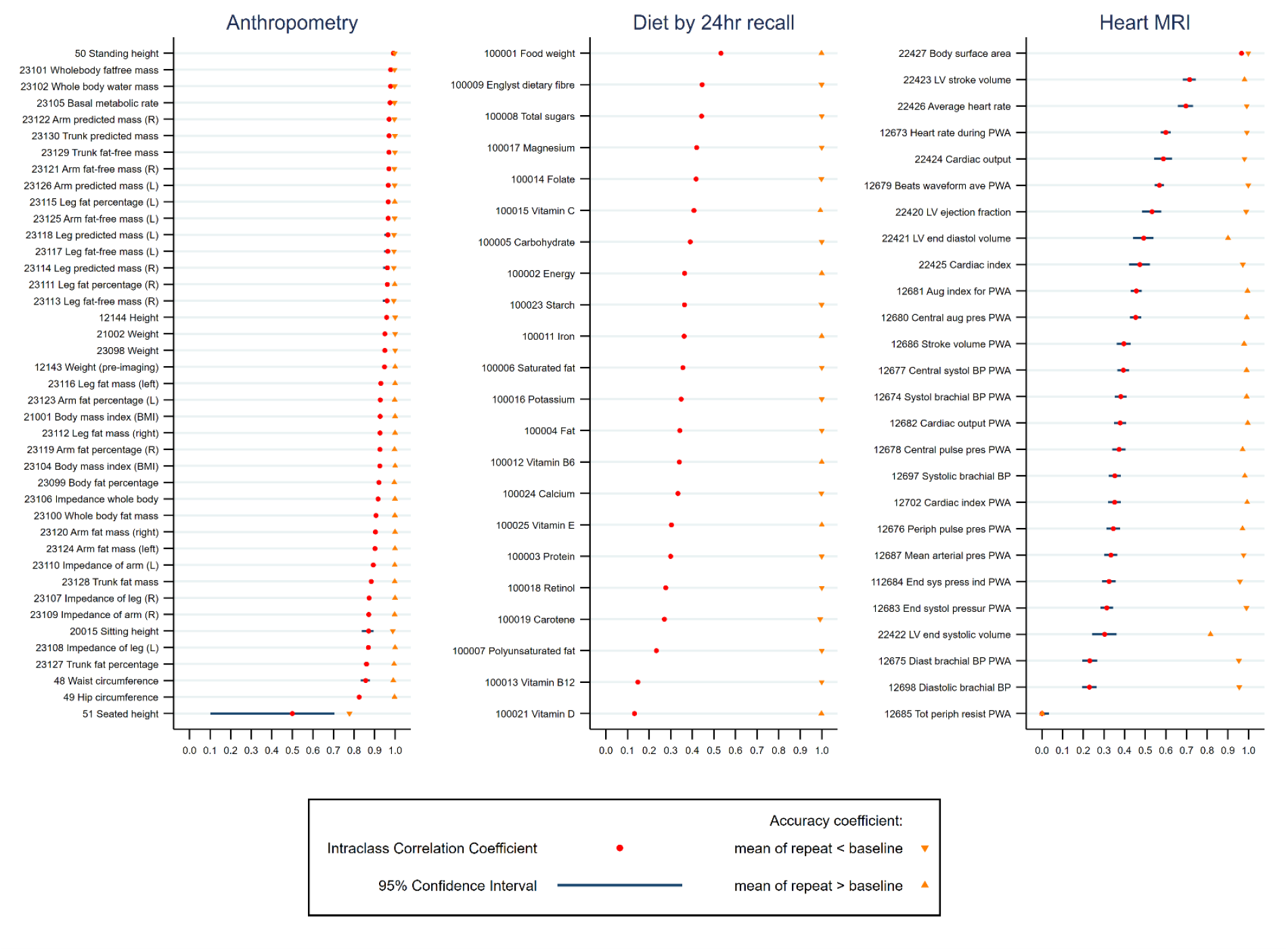
Panel of intraclass correlation coefficients for three variable categories

#### Correction for regression dilution bias – an illustrative example using CRP

The ICC for CRP suggested substantial random error (ICC=0.29, 95% CI=[0.27, 0.30]). BMI and pack-years of smoking had evidence of little random error (ICC=0.93, 95% CI=[0.92, 0.93] and ICC=0.85, 95% CI=[0.84, 0.86], respectively). The accuracy coefficients for all three were 1.00 indicating little systematic error (Table 1 and Table S4). Across 3 739 698 person years of follow up (an average of 11.8 years per participant) there were 23 021 deaths, giving an all-cause mortality rate of 6.16 per 1000 person-years (95% CI=[6.08, 6.24]); comparative rates for cardiovascular and cancer mortality were 1.12 (95% CI=[1.09, 1.16]) and 3.15 (95% CI=[3.09, 3.20]), respectively. Higher levels of CRP were associated with higher risk of all-cause, cardiovascular and cancer mortality in all analyses (Table 2). In confounder adjusted analyses for all-cause mortality with no correction for bias due to random error, the hazard ratio was 1.029 (95% CI: 1.028, 1.031) per 1mg/L higher CRP (Table 2). With correction for random error in the exposure the hazard ratio increased to 1.107 (95% CI=[1.099, 1.116]) using the ICC correction approach and to 1.119 (95% CI=[1.095, 1.143]) using the regression calibration approach. With further correction for random error in the smoking and BMI confounders there was little further change (Table 2). Patterns of change with correction for bias due to exposure and confounder random error for cardiovascular and cancer mortality were similar to those for all-cause mortality (Table 2).

**Table 1:** Measures of random and systematic error for C-reactive protein, BMI and pack years smoking for illustrative example

| UK Biobank field number | UK Biobank field name | Sample size | Accuracy coefficient<sup>a</sup> | Intraclass correlation coefficient<sup>b</sup> (95% CI) | Correction factor<sup>c</sup>, $\hat{\lambda}$ | $\text{Var}(\hat{\lambda})$ | $\text{Var}(1/\hat{\lambda})$ |
| --- | --- | --- | --- | --- | --- | --- | --- |
| 30710 | C-reactive protein | 16549 | 1.00 | 0.29 (0.27, 0.30) | 3.4987 | 0.007635 | 0.000051 |
| 21001 | Body mass index | 20257 | 1.00 | 0.93 (0.92, 0.93) | 1.0794 | 0.000001 | 0.000001 |
| 20161 | Pack years of smoking | 4744 | 1.00 | 0.85 (0.84, 0.86) | 1.1745 | 0.000030 | 0.000016 |

<sup>a</sup> Intraclass correlation coefficient is a measure of overall agreement between two measures.
<sup>b</sup> Accuracy coefficient is a measure of systematic error between two measures.
<sup>c</sup> Correction factor ( $\hat{\lambda}$ ) is the reciprocal of the intraclass correlation coefficient.

**Table 2:** Cox proportional hazard results for C-reactive protein (CRP) and all-cause, cardio vascular, and cancer mortality, adjusted for sex, age, ethnicity, BMI, smoking pack years, drinking, deprivation index (n=317 917)

| Model | All-cause mortality hazard ratio of 1mg/L increase in CRP (95% confidence interval) | Cardiovascular mortality hazard ratio of 1mg/L increase in CRP (95% confidence interval) | Cancer mortality hazard ratio of 1mg/L increase in CRP (95% confidence interval) |
| --- | --- | --- | --- |
| (i) Uncorrected Cox proportional hazard | 1.029 (1.028, 1.031) | 1.027 (1.023, 1.031) | 1.027 (1.024, 1.029) |
| (ii) Corrected for regression dilution in CRP (exposure) using intraclass correlation coefficient | 1.107 (1.099, 1.116) | 1.097 (1.081, 1.114) | 1.097 (1.087, 1.108) |
| (iii) Corrected for regression dilution in CRP using regression calibration<sup>a</sup> | 1.119 (1.095, 1.143)<sup>c</sup> | 1.108 (1.083, 1.134)<sup>c</sup> | 1.108 (1.085, 1.131)<sup>c</sup> |
| (iv) Corrected for regression dilution in CRP and both body mass index and smoking pack years (confounders) using regression calibration<sup>b</sup> | 1.118 (1.095, 1.142)<sup>c</sup> | 1.106 (1.081, 1.131)<sup>c</sup> | 1.107 (1.085, 1.130)<sup>c</sup> |

<sup>a</sup> regression calibration based on linear regression model for repeated measures of CRP (n=11 048) adjusted for all confounders
<sup>b</sup> regression calibration based on linear regression models for repeated measures of CRP (n=11 048), body mass index (n=12 529) and smoking pack years (n=4387) adjusted for CRP and all confounders
<sup>c</sup> Bootstrapped 95% confidence intervals with 10 000 replications

#### Supplementary data

Supplementary data are available at *IJE* online.

## Discussion

In this paper we provide summary ICC and accuracy coefficient data for 2858 UKB continuously measured repeatedly assessed variables. We also provide code for researchers to adapt to explore the impact of random measurement error in exposures and confounders in future UKB studies. This is important as a subgroup of participants had a repeat assessment, at cost to the funders and participants, in order to support assessment and correction for random measurement error, but to date this has been done for just a tiny fraction of the UKB studies. The provided analysis code could also be useful for researchers wanting to explore the impact of random measurement error in other studies with repeat measures.

### Assumptions of correcting for random measurement error including RDB

A key assumption when correcting for bias due to random measurement error in covariables is that this random error is non-differential to the outcome (i.e. random error in the covariable is unrelated to the observed values of the outcome). If this assumption does not hold then the association could be biased in either direction and results of adjustment could be invalid. A further assumption is that there is some true underlying or average value of the exposure of interest that is either measured with error, fluctuates regularly, or both. If a specific measurement at one particular time point is the target variable, then adjusting for RDB would not be appropriate. In our illustrative example we were interested in the effect of the underlying average value of CRP, not the value on a particular day that may change due to acute infection. Lastly, in correcting for random error in an exposure and some confounders there is an implicit assumption that no other (e.g. binary or unmeasured) confounders are also subject to random measurement error, otherwise results may still be biased in either direction. These assumptions should be carefully considered to determine the appropriateness of correcting for random error in exposures and confounders; if it is unclear whether the assumptions have been met, results adjusted for measurement error in covariables might be best considered as sensitivity analyses.

### Study strengths and limitations

The main strength of this paper lies in the large sample size and breadth of variables available in UKB. This has allowed us to explore the extent that random error varies across different categories of variables. The fact that UKB is widely used means there is the potential for this paper to help increase the number of studies correcting for random error in exposures and confounders, and the methods can also be used for other studies where repeat measures are available.

The median time between repeat measurements was greatest and most variable for the baseline assessment repeats, with consistent and shorter gaps for the online diet and image variables. With longer periods between measures, differences may reflect systematic differences. For example, the two measures with the longest time between repeats were smoking variables and measurements of antigens to different infectious diseases, each with average differences of 4.5 years. In that period of time differences in pack-years of smoking could reflect changes in behaviour due to ill health, and differences in antigen levels could reflect recent or chronic infection, rather than random variation. Thus, it is important to consider the time between measurements, as well as the nature of the measures, when considering whether the assumptions of correcting for random error (see above) might be violated.

Misclassification in categorical covariables can cause bias in the association of interest but current methods for dealing with bias due to random measurement error cannot address this (unless there are a large number of ordinal categories that can be treated as continuous). Such measurement error, along with other forms of potential bias (such as non-random error in exposure, confounders and outcome), should always be considered when interpreting results.

### Conclusions and recommendations

Greater consideration should be given to exploring bias due to random error in exposures and confounders. Investigators setting up prospective studies should be encouraged to provide repeat measures in a sub-sample for this purpose. If repeat measurements are available, then they should be used to investigate and potentially account for random error in exposures and confounders.

It is important to explore random error in all covariables (where possible) before using an adjustment. Adjusting for RDB in the exposure only will never result in a weaker association being found. However, if there is also random error in the confounders then the estimates could be biased in either direction. Where repeat measures are not available in a study of interest, it may be possible to use a correction factor from another study to correct for exposure-outcome RDB correction, but this would not allow correction for random error in confounders.

For transparency we recommend presenting all results in analyses, as in our illustrative example, including the ICC and accuracy coefficient for exposure and confounders, along with three sets of results that account for: i) no measurement error in covariables, ii) measurement error in exposure, and iii) measurement error in exposure and confounders.

## Data Availability

All data produced in the present work are contained in the manuscript or supplement. Underlying data used is available by application to UK Biobank. Code used is available on GitHub.

https://github.com/MRCIEU/measurement_error_adjustment

## Ethics approval

UKB received ethical approval from the UK National Health Service’s National Research Ethics Service [ref 11/NW/0382].

## Acknowledgments

This research was conducted using the UKB Resource under Application number 16729 [dataset IDs 48196 and 41913]. We are thankful to all participants for giving their time and data as well as to all those who created and maintain this open access resource for researchers. This work used the computational facilities of the Advanced Computing Research Centre, University of Bristol - http://www.bristol.ac.uk/acrc/.

## Author contributions

CER, DAL, MCB, LACM designed the analytical strategy. CER ran the analysis and visualization, and wrote the original draft. DAL, MCB and LACM reviewed and edited the draft manuscript.

