## Supplementary text and figures for "Exploring regression dilution bias using repeat measurements of 2858 variables in up to 49 000 UK Biobank participants"

**Supplementary Material**

### Text S1. Data preparation details

UKB assigns specific default values to some numeric variables to denote non-numeric answers or missing *values* (e.g. 'unknown' or 'less than a mile'). We replaced these with an appropriate numerical value where possible (e.g. 0.5 for 'less than a mile') or else classed it as missing data. Details are in supplementary Table S2.

In addition, some variables include multiple readings for an individual at a single visit, for example blood pressure was measured twice at each visit. We combined such multiple measurements in an appropriate way to create one analysis variable, for example by taking the mean. Details are in Supplementary Table S3.

### Text S2. Formulae for the intraclass correlation coefficient (ICC) and the accuracy coefficient

The Stata command for the two-way mixed effects ICC for absolute agreement, used with long format data is: **icc *variable id measure*, mixed absolute** where *variable* is the name of the variable of interest, *id* is the variable containing the participant id, *measure* is an indicator variable for the measurement type (initial or repeat).

The formula for this ICC is: $$\frac{MS_I - MS_E}{MS_I + \frac{MS_M - MS_E}{n}}$$

where $MS_I$=mean square for individuals, $MS_E$=mean square for error, $MS_T$=mean square for measures

From *Koo TK, Li MY. A Guideline of Selecting and Reporting Intraclass Correlation Coefficients for Reliability Research. J Chiropr Med. 2016 Jun;15(2):155-63*

The accuracy coefficient is a component of Lin's Concordance Correlation Coefficient (CCC). The CCC can be partitioned into Pearson's correlation coefficient and the accuracy coefficient as follows:

$$\text{CCC} = \frac{2s_{xy}}{s_y^2 + s_x^2 + (\bar{y} - \bar{x})^2}$$

$$= \frac{s_{xy}}{s_x s_y} * \frac{2 s_x s_y}{s_y^2 + s_x^2 + (\bar{y} - \bar{x})^2}$$

Pearson's correlation coefficient $= \frac{s_{xy}}{s_x s_y} \qquad \in [0,1]$

Accuracy coefficient $= \frac{2 s_x s_y}{s_y^2 + s_x^2 + (\bar{y} - \bar{x})^2} \qquad \in [0,1]$

where $s_x$ is the sample standard deviation of the baseline values, $s_y$ is the sample standard deviation of the repeat values, $s_{xy}$ is the sample covariance of the two measurements, $\bar{x}$ is the sample mean of the baseline values and $\bar{y}$ is the sample mean of the repeat values.

From *Lin L., A Concordance Correlation Coefficient to Evaluate Reproducibility, 1989 Biometrics 45, 255-268*

### Text S3. Variance formulae for the correction factor

The formulae for the variance of the estimated ICC-based correction factor ($\hat{\lambda}$) and the variance of its reciprocal ($1/\hat{\lambda}$) are as follows:

$$Var(\hat{\lambda}) \approx \frac{(\hat{\lambda}^2 - 1)^2}{n}$$

$$Var\left(\frac{1}{\hat{\lambda}}\right) \approx \frac{Var(\hat{\lambda})}{\hat{\lambda}^4}$$

where n is the number of individuals with repeat measurements.

From *Frost C, Thompson SG. Correcting for regression dilution bias: comparison of methods for a single predictor variable. J R Statist Soc A 2000; 163(2)173-89*

### Text S4. Confounder definitions for illustrative example

Sex is a baseline binary field in UKB (field 31).

Age at recruitment is a continuous variable (UKB field 21022) included as a linear covariate.

Deprivation score was made by linking the 3 UKB Indexes of Multiple Deprivation for separate home nations together (fields 26410, 26427 and 26426, only one in use for each individual).

Ethnic background is a categorical UKB variable (field 21000), used at the top level with 6 categories of white, mixed, Asian/Asian British, black/black British, Chinese, and other.

Drinking uses the UKB self-reported Alcohol intake frequency question (field 1558) with categories of daily or almost daily, 3 or 4 times a week, Once or twice a week, 1-3 times a month, special occasions only, and never.

BMI is a continuous variable in UKB (field 21001) that is used as a linear covariate.

Smoking is a calculated continuous variable based on Pack years of smoking (field 20161, for people who have ever smoked over the age of 16), the binary variable Ever smoked (field 20160), Age stopped smoking (field 22507, used to exclude those who stopped before 16 from being classed as smokers). The definition in Stata code is as follows:

```
gen smoking=smokpack
replace smoking=0 if smokever==0 & smokpack==.
replace smoking=0 if smokever`i'==1 & smokagestop`i'>0 & smokagestop`i'<=16 & smokpack`i'==.
replace smoking=. if smokever`i'==1 & smokagestop`i'<=0 & smokpack`i'==.
replace smoking=. if smokever`i'==1 & smokagestop`i'>16 & smokagestop`i'<. & smokpack`i'==.
replace smoking=. if smokever`i'==1 & smokagestop`i'==. & smokpack`i'==.
replace smoking=. if smokever==. & smokpack==.
```

Figure S1. Intraclass correlation coefficient and accuracy coefficient (from Lin's Concordance correlation coefficient) for anthropometry measures

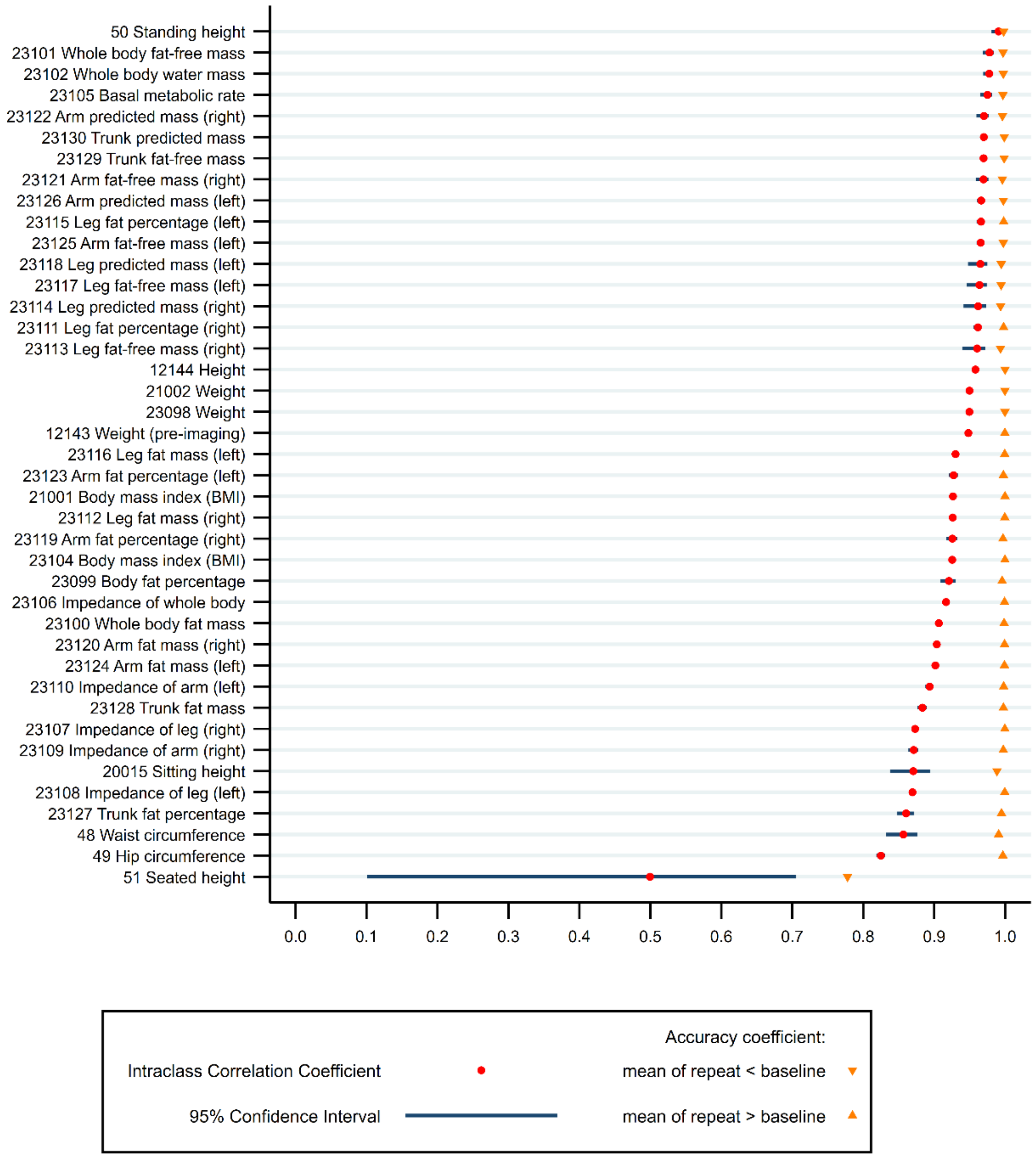

Figure S2. Intraclass correlation coefficient and accuracy coefficient (from Lin's Concordance correlation coefficient) for eye measures

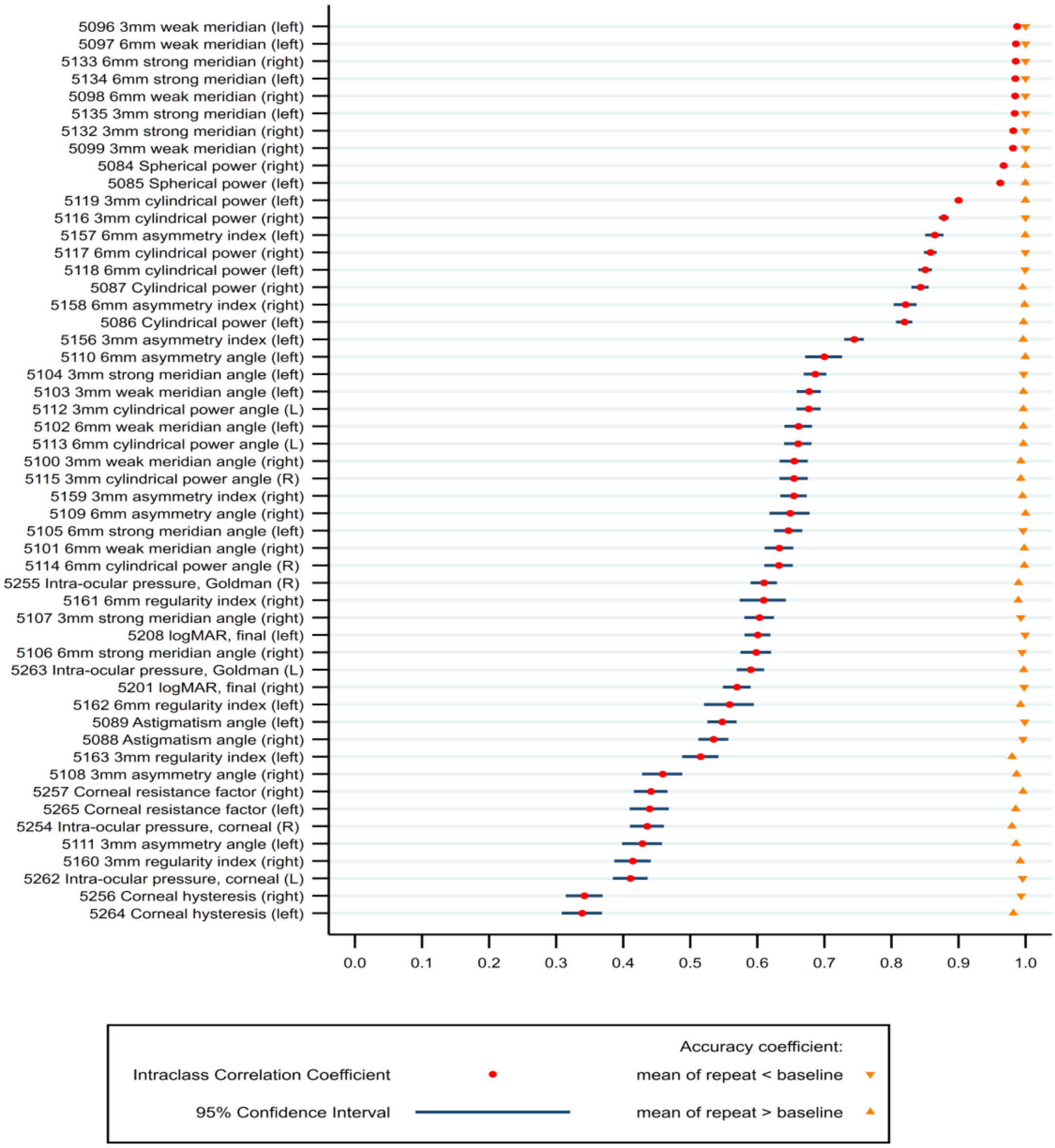

Figure S3. Intraclass correlation coefficient and accuracy coefficient (from Lin's Concordance correlation coefficient) for physical measures

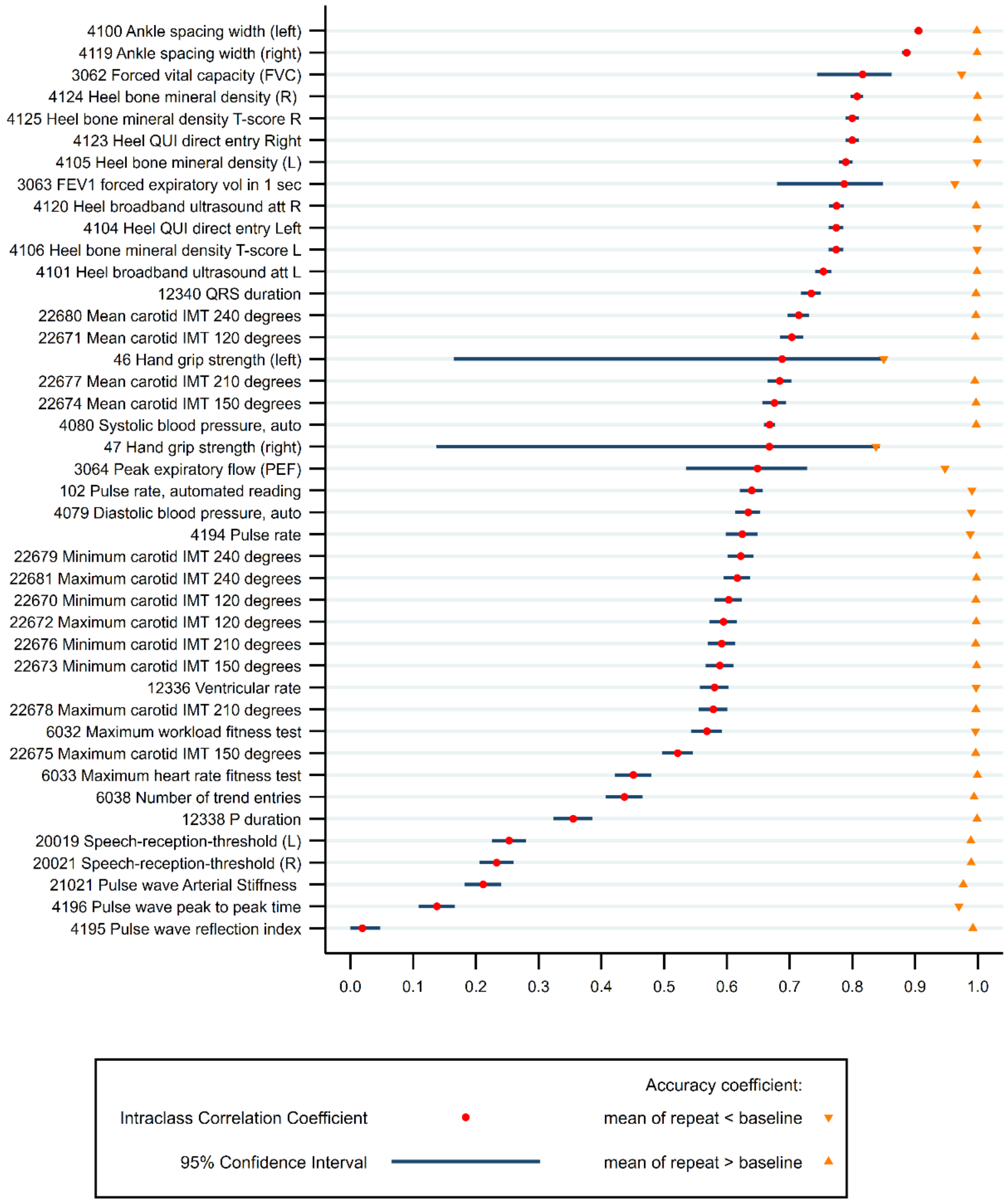

Figure S4. Intraclass correlation coefficient and accuracy coefficient (from Lin's Concordance correlation coefficient) for blood and urine biomarkers

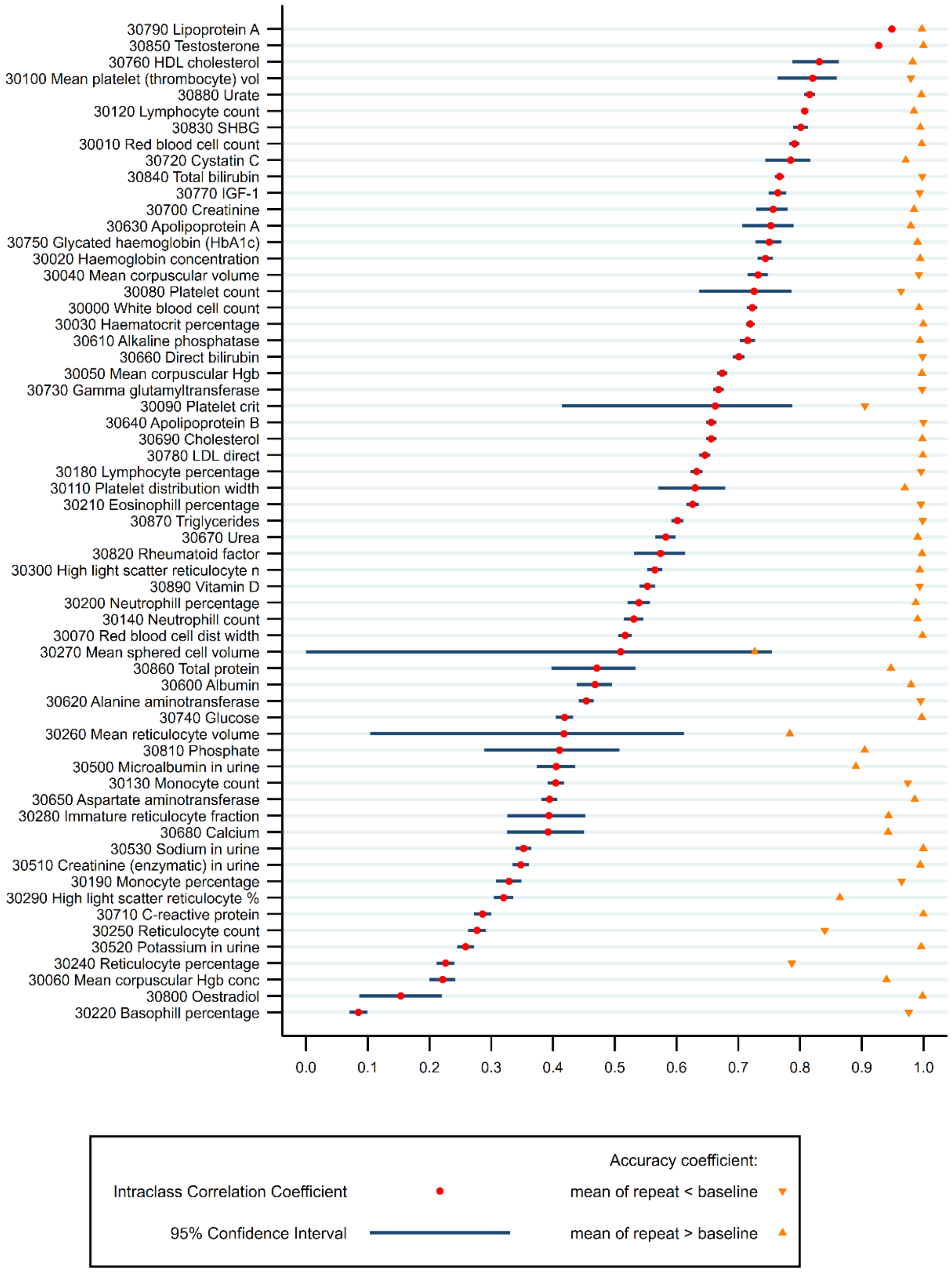

Figure S5. Intraclass correlation coefficient and accuracy coefficient (from Lin's Concordance correlation coefficient) for infectious disease antigens in blood

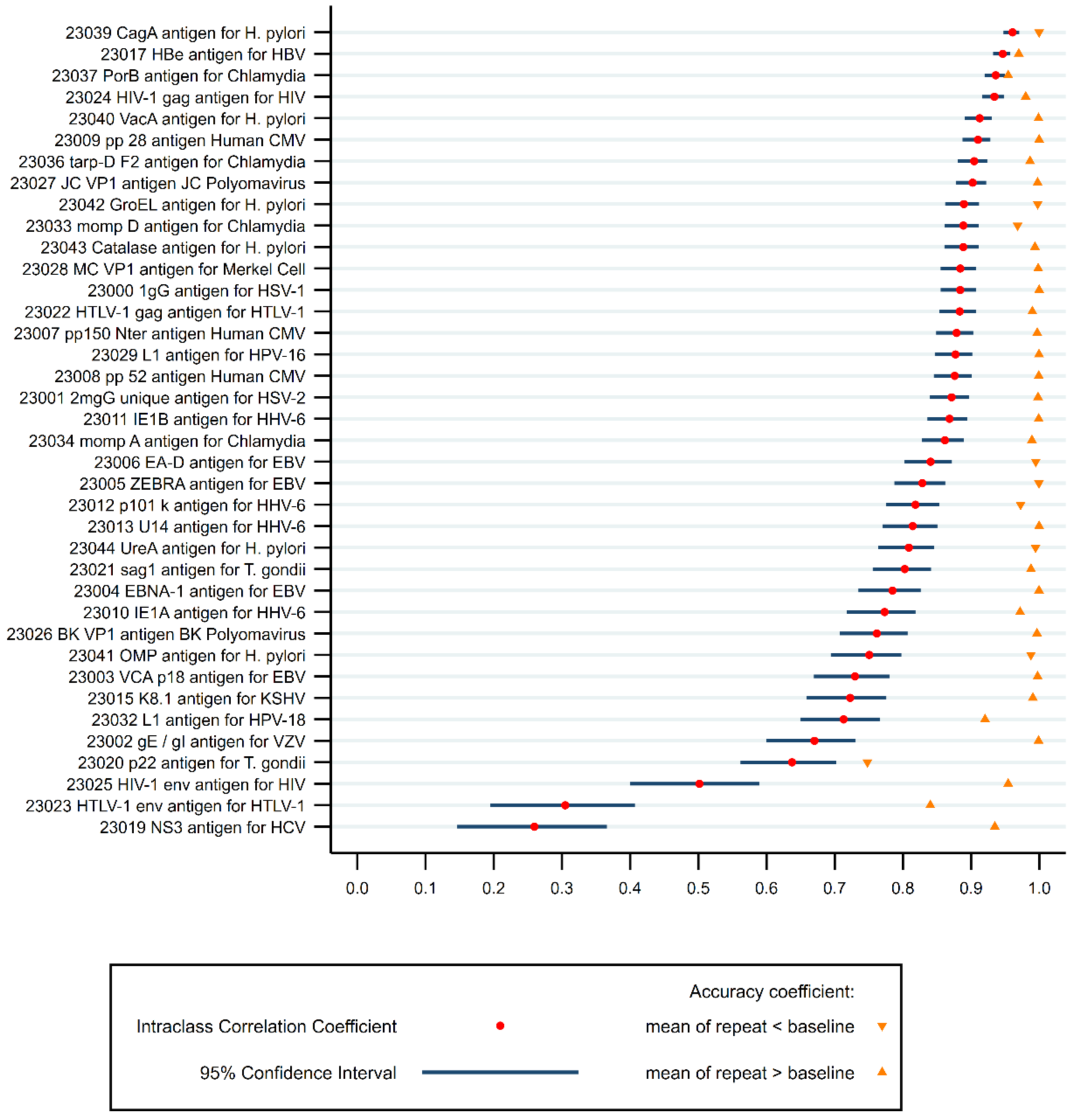

Figure S6. Intraclass correlation coefficient and accuracy coefficient (from Lin's Concordance correlation coefficient) for diet by 24hr recall measures

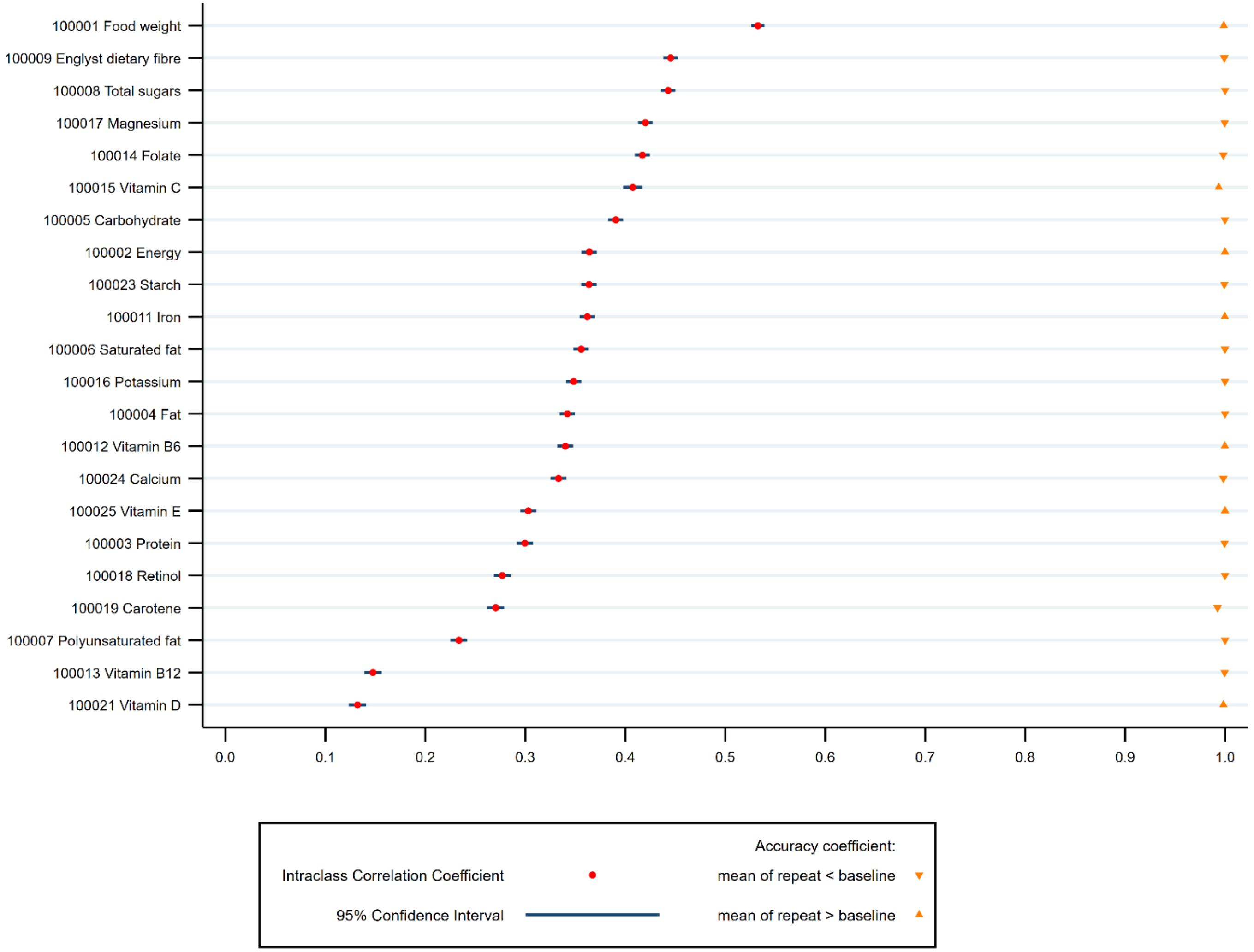

Figure S7. Intraclass correlation coefficient and accuracy coefficient (from Lin's Concordance correlation coefficient) for medical history information

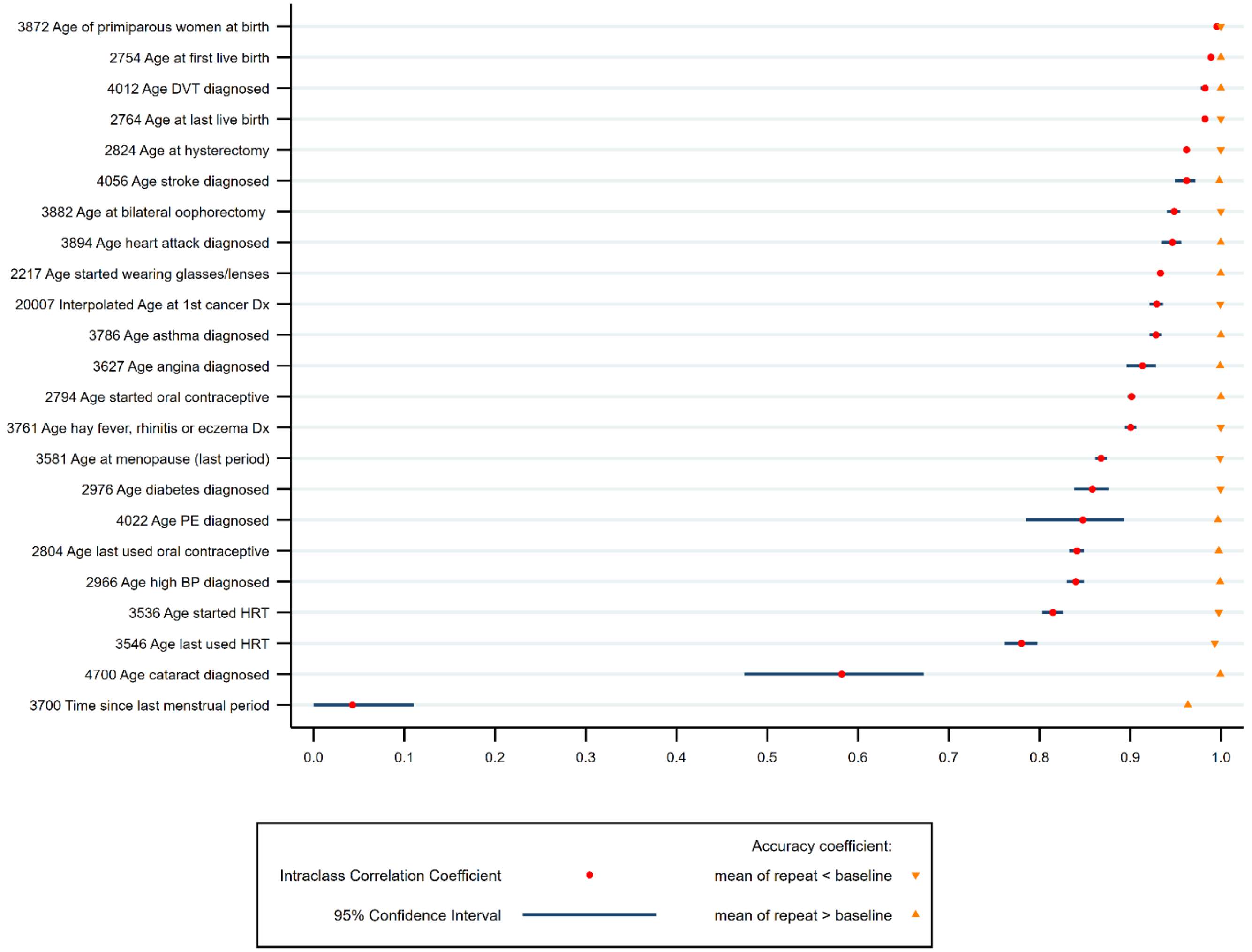

Figure S8. Intraclass correlation coefficient and accuracy coefficient (from Lin's Concordance correlation coefficient) for lifestyle measures

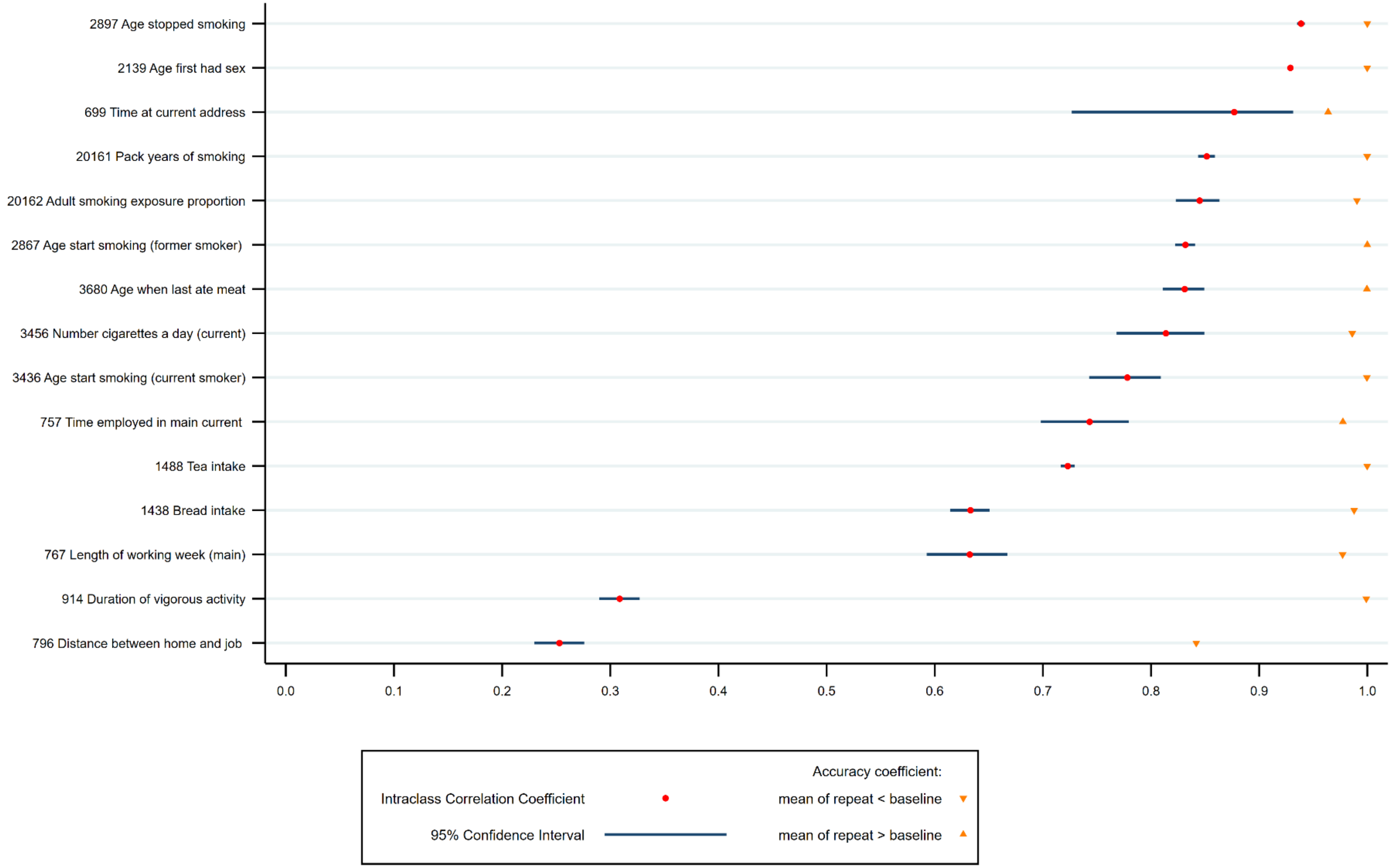

Figure S9. Intraclass correlation coefficient and accuracy coefficient (from Lin's Concordance correlation coefficient) for heart MRI measures

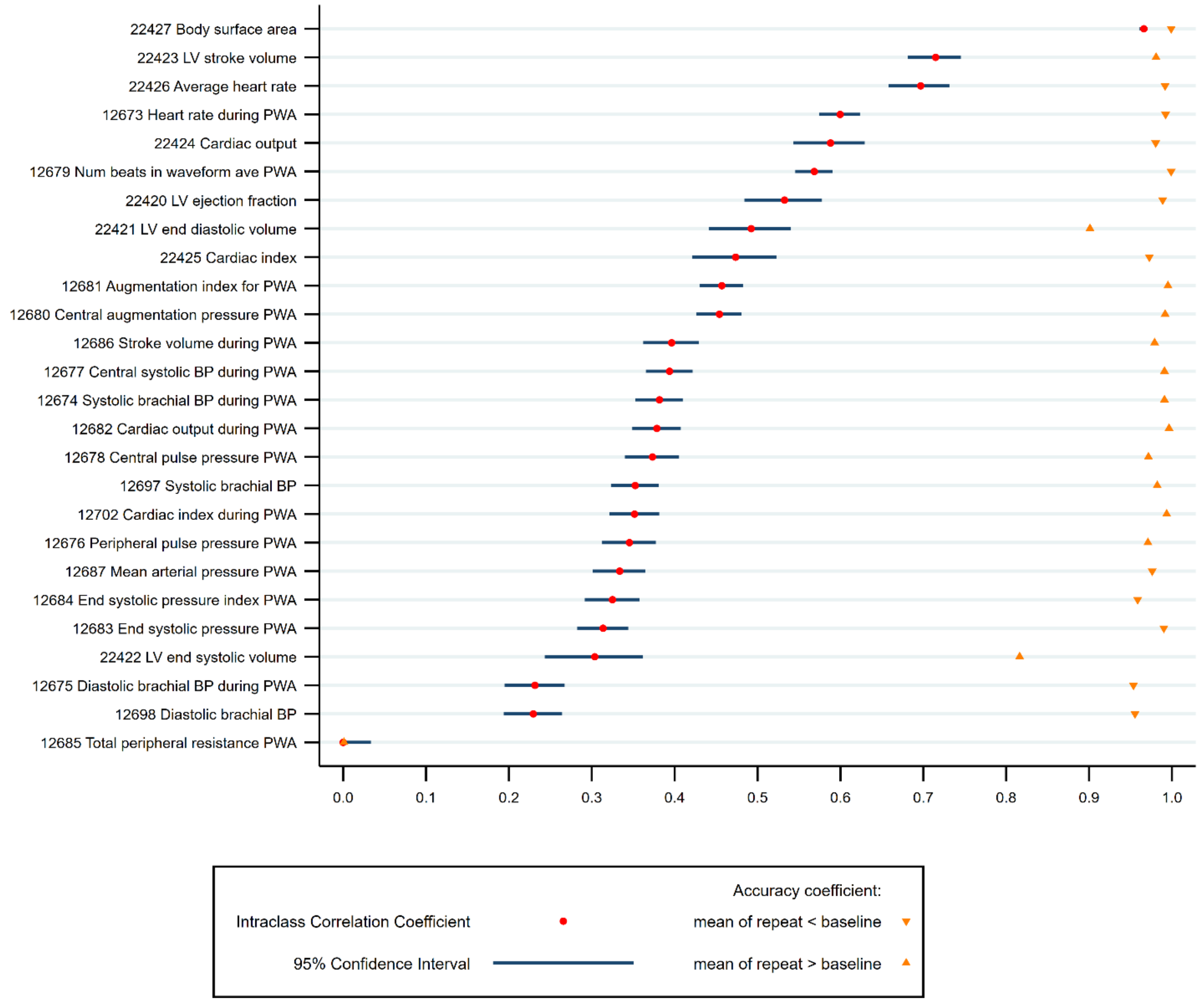

Figure S10. Intraclass correlation coefficient and accuracy coefficient (from Lin's Concordance correlation coefficient) for other measures

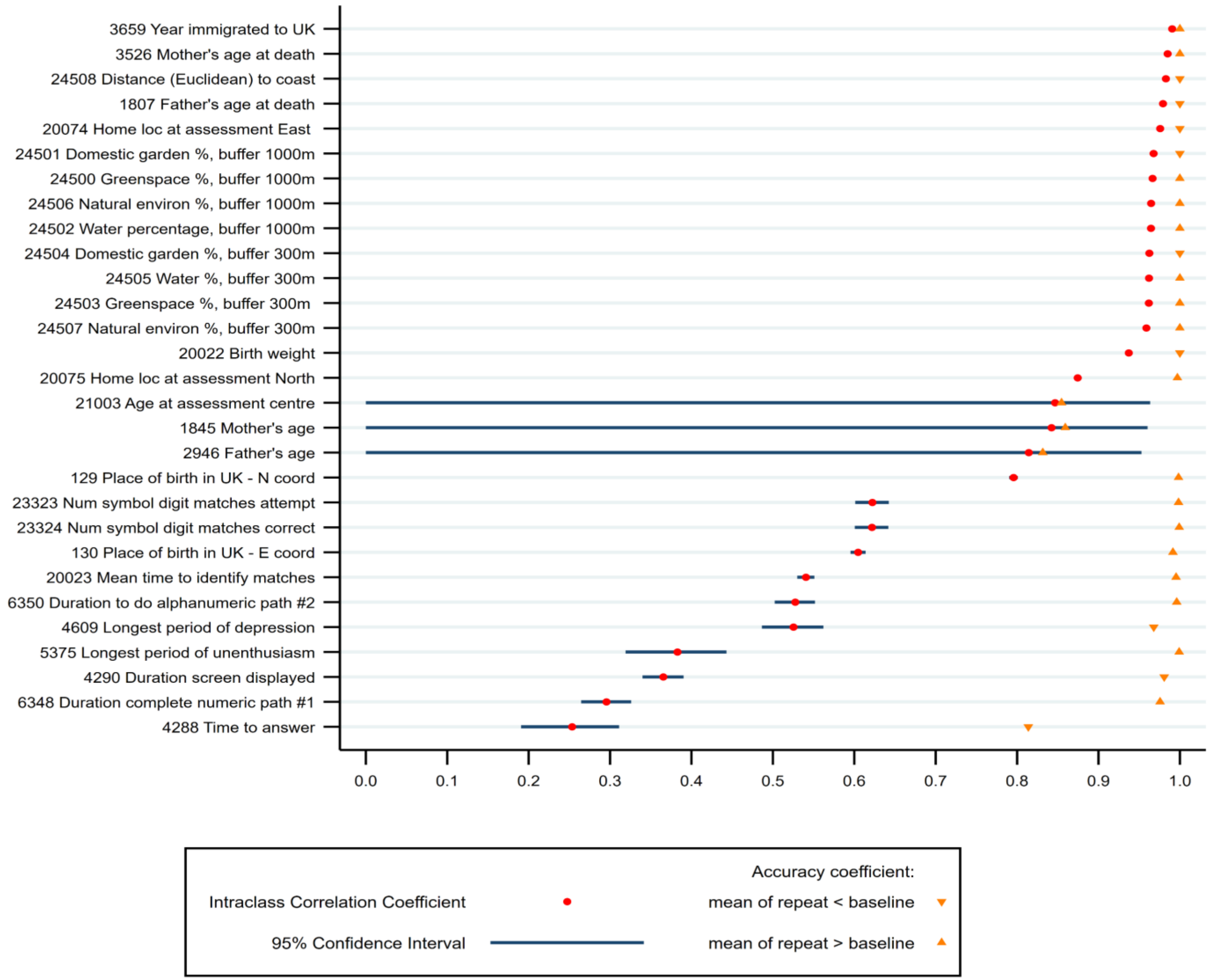
